# Dietary Meat, Plasma Metabolites, and Cardiovascular Disease Risk: A Multi-Cohort Study in Sweden

**DOI:** 10.1101/2024.10.21.24315788

**Authors:** Getachew Arage, Koen F. Dekkers, Luka Marko Rašo, Ulf Hammar, Ulrika Ericson, Susanna C Larsson, Hanna Engel, Gabriel Baldanzi, Kamalita Pertiwi, Sergi Sayols-Baixeras, Rikard Landberg, Johan Sundström, J Gustav Smith, Gunnar Engström, Johan Ärnlöv, Marju Orho-Melander, Lars Lind, Tove Fall, Shafqat Ahmad

**Author notes:** Corresponding: Dr Shafqat Ahmad, School of Natural Sciences, Technology and Environmental Studies, Södertörn University Sweden.

## Abstract

**Background:** Higher meat intake has been associated with adverse health outcomes, including cardiovascular disease (CVD). However, the mechanisms by which meat consumption increases CVD risk remain unclear. We used metabolomics data from a large population-based study to identify plasma metabolites associated with self-reported meat intake and associations with cardiometabolic biomarkers, subclinical CVD markers and incident CVD.

**Methods:** We investigated the association between self-reported meat intake and 1272 plasma metabolites measured using ultra-high-performance liquid chromatography coupled with mass spectrometry in the SCAPIS (n=8,819; aged 50-64) cohort. Meat-associated metabolites were further analyzed in relation with subclinical CVD markers in the POEM cohort (n=502, all aged 50) and with incident CVD in the EpiHealth cohort (n=2,278; aged 45-75; 107 incident cases over 9.6 years follow-up). Meat intake was assessed through food frequency questionnaire, and categorized into white, unprocessed red, and processed red meat. We analyzed associations between meat intake and metabolites, meat-associated metabolites with cardiometabolic biomarkers, and subclinical CVD markers employing linear regression, adjusting for demographics and lifestyle factors. Cox proportional hazards analysis evaluated the associations between meat-associated metabolites and CVD incident.

**Results:** After correction for multiple testing, we identified 458, 368, and 403 metabolites associated with self-reported white, unprocessed red and processed red meat intake, respectively. Metabolites positively associated with all three meat types were related with higher plasma levels of apolipoprotein A1, C-reactive protein, and increased intima-media thickness, while metabolites negatively associated were related with higher fasting insulin levels. Processed red meat-associated metabolites were related with higher levels of fasting insulin, glycated hemoglobin, and lipoprotein(a) and were inversely related with maximal oxygen consumption. Two metabolites, 1-palmitoyl-2-linoleoyl-GPE (16:0/18:2) (HR: 1.32; 95% CI: 1.08, 1.62) and glutamine degradant (HR: 1.35; 95% CI: 1.07, 1.72), associated with higher intakes of all three meat types were also related with a higher risk of incident CVD.

**Conclusion:** This study identified hundreds of metabolites associated with self-reported intake of different meat types. Processed red meat increasing metabolites were associated with worse glycemic measures and reduced cardiovascular function. These findings may enhance our understanding of the relationship between meat intake and CVD, providing insights into underlying mechanisms.

**What is New?:**

- Our study provides the most comprehensive analyses of self-reported meat intake and plasma metabolites, identifying hundreds of meat-associated metabolites using a large-scale epidemiological sample.
- We uncovered novel metabolites associated with white, unprocessed, and processed red meat intake and their association with subclinical markers and incident CVD.

**What Are the Clinical Implications?:** Meat-associated metabolites and their relationships with cardiometabolic biomarkers, subclinical markers, and CVD incident may highlight metabolic responses to meat intake and their potential impact on cardiometabolic health and CVD risk.

---

Meat is an important dietary component as it provides essential nutrients including protein and micronutrients such as iron, zinc and vitamin B12^1^. Nevertheless, epidemiological studies have shown that a high intake of dietary meat associates with an increased risk of cardiovascular disease (CVD) incident^2,3^ and its risk factors, including blood lipids, insulin, glucose, HbA1C, and C-reactive protein. However, significant variations in these associations have been observed across studies^4,5^. The mechanisms by which meat intake may influence CVD risk are complex. Heme iron content in red meat, along with saturated fats, processing methods involving sodium and nitrates which are precursors of nitrosamines, and high temperature heating, contribute to advanced glycation- and other Maillard products, which may contribute to the development of CVD^6,7^. White meat, such as chicken and turkey, is a lower-fat and lower-iron alternative to red meat and may protect against CVD, although evidence is conflicting^8^. Thus, understanding the mechanisms through which meat intake may increase the risk of CVD remains challenging and requires further exploration.

Plasma metabolites profiling presents a valuable approach for investigating micromolecules associated with habitual meat intake providing insights into its potential association with CVD^9,10^. High-throughput metabolomics offers a reliable method for assessing dietary intake, enabling a more comprehensive understanding of the relationship between meat consumption and CVD risk, particularly in the presence of heterogeneity in the study results^11^.

Previous studies^10,12–19^ were limited by small sample sizes, highlighting the need for large-scale epidemiological studies to provide robust evidence of associations between specific meat intake and related metabolites. While previous studies suggest an association between meat intake and plasma metabolites, investigating the relationship between meat-associated metabolites and cardiometabolic biomarkers, subclinical CVD markers, or CVD incident remains limited^20^.

Therefore, our primary aim was to investigate the association between self-reported white meat, unprocessed red meat, and processed red meat intake with plasma metabolites using the Swedish CArdioPulmonary bioImage Study (SCAPIS) cohort. As a secondary aim, we further investigated the identified meat-associated metabolites in relation to cardiometabolic biomarkers in the SCAPIS cohort, their relationship with subclinical CVD markers in the POEM cohort, and incident CVD in the EpiHealth cohort.

## Methods

### Study population

The current study included data and samples from three population-based studies: SCAPIS, POEM, and EpiHealth. The studies were conducted in compliance with the principles of the Declaration of Helsinki and were approved by the responsible ethics committees (DNR 2023-07352-01, DNR 2009-057, DNR 2018-315), and written informed consent was obtained from all participants.

The SCAPIS is a population-based study conducted in Sweden between 2013 and 2018, involving 30154 men and women aged 50-64 years ^21^. For the current analyses, we used two subcohorts of SCAPIS, namely SCAPIS-Uppsala (n=5,036) and SCAPIS-Malmö (n=6,251). In SCAPIS, metabolomic measurements were available for 8,962 participants^22^. All participants attended the study sites for a minimum of two visits. During visit 1, participants completed a questionnaire, including a food-frequency questionnaire (FFQ) and self-reported history of morbidities and medications. Participants also underwent anthropometric measurements and provided venous blood samples. In visit 2, blood pressure measurement was performed. The average duration between visits 1 and 2 was 15 days in Uppsala and 11 days in Malmö. The flowchart of the study design is shown in the supplementary figure (**Figure S1)**.

The POEM study was conducted in men and women living in Uppsala, Sweden, all aged 50 years, from 2010 to 2016 (n=502)^23^. In this study, the subclinical markers of CVD included intima-media thickness of the carotid artery (IMT), intima-media gray-scale median (IMGSM), fasting augmentation index (AIx), endothelium-dependent vasodilation (EDV), endothelium-independent vasodilation (EIDV), flow-mediated vasodilation (FMD), maximal oxygen consumption (VO2max), hyperemia brachial artery blood flow (HBABF), resting brachial artery blood flow (BABF), resting forearm blood flow (FBF) and pulse wave velocity (PWV). The assessment of these subclinical markers of CVD in POEM is described in detail in the extended methods in the supplementary material.

For our analysis of meat-associated metabolites in relation to incident CVD, we utilized the published summary results from EpiHealth^24^. The EpiHealth was conducted in Uppsala and Malmö from 2011 to 2018, and metabolomics data was available exclusively from the Uppsala segment (n=2,278). Participants were followed up for an average of 9.6 years after metabolomics measurement, using data from Swedish registries of mortality and in-hospital care. During this period, 107 incident cases of CVD were identified. The combined endpoint included myocardial infarction (ICD-10 code I21), ischemic stroke (ICD-10 code I63), and heart failure (ICD-10 codes I50 and I11.0), covering both fatal and nonfatal events.

### Meat intake assessment

Meat intake was assessed using a web-based FFQ (MiniMeal-Q), previously validated for the SCAPIS cohort^25,26^. Briefly, the FFQ was self-administered, semi-quantitative, and included questions about diet and portion sizes. The FFQ covered 75-126 commonly consumed food items in Sweden. During their initial visit, participants were asked to indicate how often, on average, they had consumed various foods for a month, ranging from five times a day to one-to-three times a month. In this study, meat intake was categorized into white meat, unprocessed red meat, and processed red meat, based on classifications made in previous Swedish studies^27,28^. White meat included chicken/other poultry; unprocessed red meat included pork, beef, lamb, game, hamburger, kebab, and minced meat dishes; and processed red meat included ham/salami for sandwiches and sausages. Meat intake (g/d) was calculated using frequency of meat intake and portion size information for all three meat types.

### Plasma metabolome profiling

In the three datasets, the same untargeted metabolomics measurement (Metabolon) was performed on plasma samples, ensuring that the identified metabolites were consistent across all studies. Fasting plasma samples were collected after overnight fasting and stored at -80^0^C in a central biobank until sent to Metabolon Inc. (Durham, NC, USA) for non-targeted metabolome analysis. The quality control standards for metabolome analysis were described in previous studies^22,24,29^.

Briefly, four modes were used to maximize metabolite coverage and identification: two separate reverse phase ultra-high-performance liquid chromatography-tandem mass spectrometry (UPLC-MS/MS) with positive ion mode electrospray ionization (ESI), reverse phase UPLC-MS/MS with negative ion mode ESI and hydrophilic interaction UPLC-MS/MS with negative ion mode ESI were used. During the annotation process, two types of metabolic pathways were assigned. The first is metabolite class, which encompasses broad pathway categories and primarily includes endogenous compounds such as amino acids and lipids. The second is metabolite subclass, which refers to more specific pathway categories. Metabolite measurements that fell below the detection threshold were imputed with the lowest observed value for each respective metabolite. In the SCAPIS study, 1,308 metabolites were included in the analysis. Metabolites having at least 100 measurements above the detection threshold resulted in a total of 1,272 metabolites included in the analysis. In this study sample, 8,957 of 8,962 participants (SCAPIS-Uppsala, n=4979; SCAPIS-Malmö, n=3978) with high-quality plasma metabolomic data were included. This is because two samples were lost during the process, as has been mentioned previously^22^. In POEM study, 1,208 out of 1,255 metabolites were analyzed, excluding those metabolites with at least 100 measurements below the detection threshold. Metabolomics measurements were conducted in 501 participants, with one sample excluded due to missing metabolite data.

### Cardiometabolic biomarker measurement

Total cholesterol, high-density lipoprotein cholesterol (HDL-C), and triglycerides were measured using standard laboratory methods. Low-density lipoprotein cholesterol (LDL-C) was calculated via the Friedewald formula^30^. Fasting plasma samples were analyzed on the Alinity C system for apolipoprotein A1 (ApoA1), apolipoprotein B (ApoB), and lipoprotein(a) (Lpa), while insulin was assessed using the Alinity I kit. Plasma glucose was determined by the hexokinase method. Hemoglobin A1c (HbA1c) was measured in SCAPIS-Uppsala using capillary electrophoresis and in SCAPIS-Malmö via turbidimetry. C-reactive protein (CRP) was measured using the immunoturbidimetric method. Blood pressure was measured using an Omron M10-IT device.

### Covariates

Self-reported covariates included age at baseline (continuous measure), sex (male/female), education level (categorized as not completed elementary school, elementary school, high school, or university/college degree), smoking status (regular/not regular), and physical activity level over the last three months (never, not regularly, 1-2 times/week, 2-3 times/week, or more than 3 times/week). Additionally, self-reported history of hypertension, medication use for hypertension, cholesterol, and/or diabetes in the last two weeks (yes/no), total energy intake (kcal per day), fibre intake, alcohol intake, and intake of fruits, vegetables, and whole grains (all grams per day) were collected from questionnaires. Body mass index (BMI) was calculated by dividing the weight in kilograms by the height in meters squared. Participants with implausible total energy intake, defined as exceeding 5000 calories for women and 6000 calories for men, or below 500 calories for women and 550 calories for men, were excluded.In the POEM study, the covariates included: sex (male/female), BMI (kg/m^2^), current smoking status, waist-hip ratio (measured as waist circumference in cm and hip circumference in cm), manually measured systolic blood pressure (SBP) and diastolic blood pressure (DBP) (mmHg), fasting blood glucose (mmol/L), creatinine, serum cholesterol (mmol/L), HDL-C (mmol/L), self-reported insulin therapy, diabetes, cholesterol-lowering medications, medications for diabetes, other lipid agents, and a history of stroke or myocardial infarction.

Moreover, summary results data about meat-associated metabolites in relation to incident CVD in the EpiHealth study was extracted from the previously published study^24^, where analyses were adjusted for covariates including age, sex, and traditional CVD risk factors such as systolic blood pressure (SBP), diabetes, LDL-C, HDL-C, BMI, and current smoking status.

### Statistical analysis

Individual-level data from the SCAPIS-Uppsala and SCAPIS-Malmö cohorts were combined for analysis. The data was checked for missing values, outliers, and statistical distributions. Missing values were addressed using complete case analysis.The characteristics of study participants were presented based on their tertiles of meat intake, using means and standard deviations (SD) for continuous variables and proportions for categorical variables. For the descriptive table, the measures of white meat, unprocessed red meat, and processed red meat intake were combined into one variable as total meat intake, which was divided into tertiles. Pearson correlation coefficients were calculated to examine the relationships between the intake of these three meat types. Plasma metabolites were log-transformed and standardized to a mean of 0 and SD of 1. The biomarkers TG, CRP, Lpa and insulin had skewed distributions, and thus were log-transformed prior to analysis. We analyzed association of meat types with metabolites using linear regression analysis. All estimates from the linear regression analyses were reported per 20-gram-per-day increase in meat intake, which corresponds to approximately a 1 SD increase for white and processed red meat, and a 0.5 SD increase for unprocessed red meat. Two sets of regression analyses were performed. First, the basic model adjusted for age, sex, metabolomic delivery batch, total energy intake, intake of fiber, whole grain, fruits and vegetables, alcohol, along with physical activity, education, smoking status, history of hypertension, and medication for hypertension, cholesterol and diabetes. Second, the full model included the same covariates as the basic model, with additional adjustment for BMI. In both models, *p*-values were corrected for multiple testing using the method proposed by Benjamini and Hochberg^31^. False discovery rate–adjusted *p*-values (*q*-values) were estimated, with the threshold set at 0.05.

Linear regression models were used to analyze the association of top 20 meat-associated metabolites with cardiometabolic biomarkers adjusted for age, sex, metabolomic delivery batch, total energy intake, alcohol intake, smoking, physical activity and education. Similarly, top 20 meat-associated metabolites in reation to subclinical CVD markers were examined using linear regression, adjusted for sex, BMI, waist-hip ratio, smoking status, SBP and DBP, fasting blood glucose, creatinine level, total cholesterol, HDL-C, insulin, history of diabetes, cholesterol-lowering drugs, medication for diabetes and other lipid agents. Furthermore, summary results about metabolite-incident CVD were extracted from the previously published finding for the Epihealth study, with details on statistical analysis and confounding variables previously reported^24^.

Enrichment analysis for metabolic pathways was performed based on ranked association *q*-values of the significant metabolites from fully adjusted model results of each meat type and metabolites association using the fast gene set enrichment analysis (fgsea) package^32^ v1.19.2 package for both positively and negatively associated metabolites. Statistical analyses were performed using Stata version 18.0 and visualizations were created using ComplexHeatmap package in R version 4.3.2^33^.

## Results

### Participant characteristics

Characteristics of the study participants are presented as the extreme tertiles (tertile 1 vs tertile 3) of total meat intake (**Table 1)**. Men reported a higher total meat intake compared to women in both SCAPIS-Uppsala and SCAPIS-Malmö. Moreover, we observed a higher mean (SD) of BMI, total energy intake, and alcohol intake among participants with higher total meat intake at both study sites. However, higher intake of fruits and vegetables, and higher concentration of HDL-C and insulin were observed among the participants in SCAPIS-Malmö. **Tables S1, S2, and S3** in the Supporting information provide the characteristics of the study participants based on the extreme tertiles of white meat, unprocessed red meat, and processed red meat intake, respectively. Pearson correlation coefficients were calculated to examine the relationships between the intake of the three meat types: white meat intake was positively correlated with unprocessed red meat intake (r=0.35, *p*-value<0.0001) and processed red meat intake (r=0.12, *p*-value<0.0001), and unprocessed red meat intake was also positively correlated with processed red meat intake (r=0.38, *p*-value<0.0001).

**Table 1.**
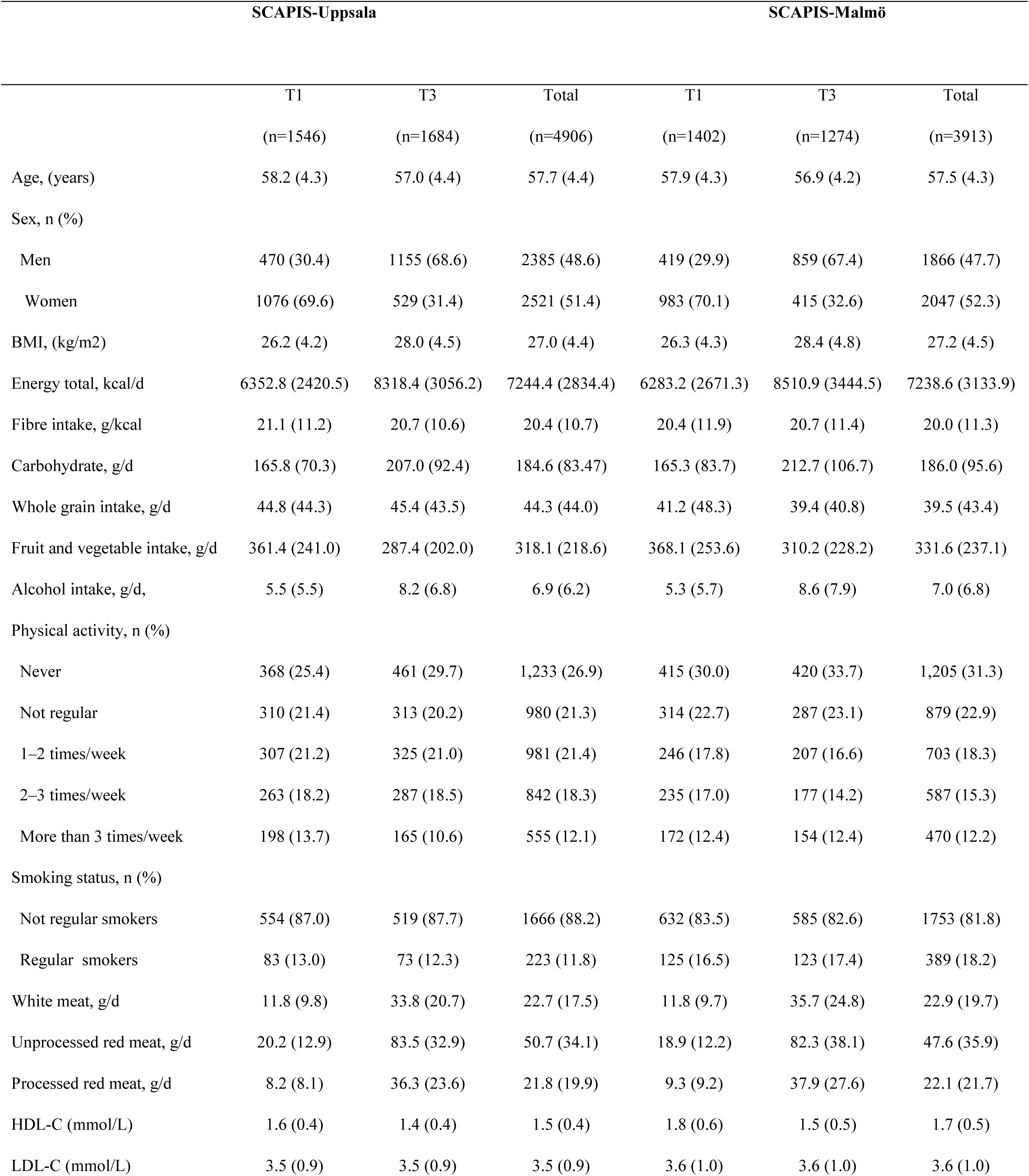
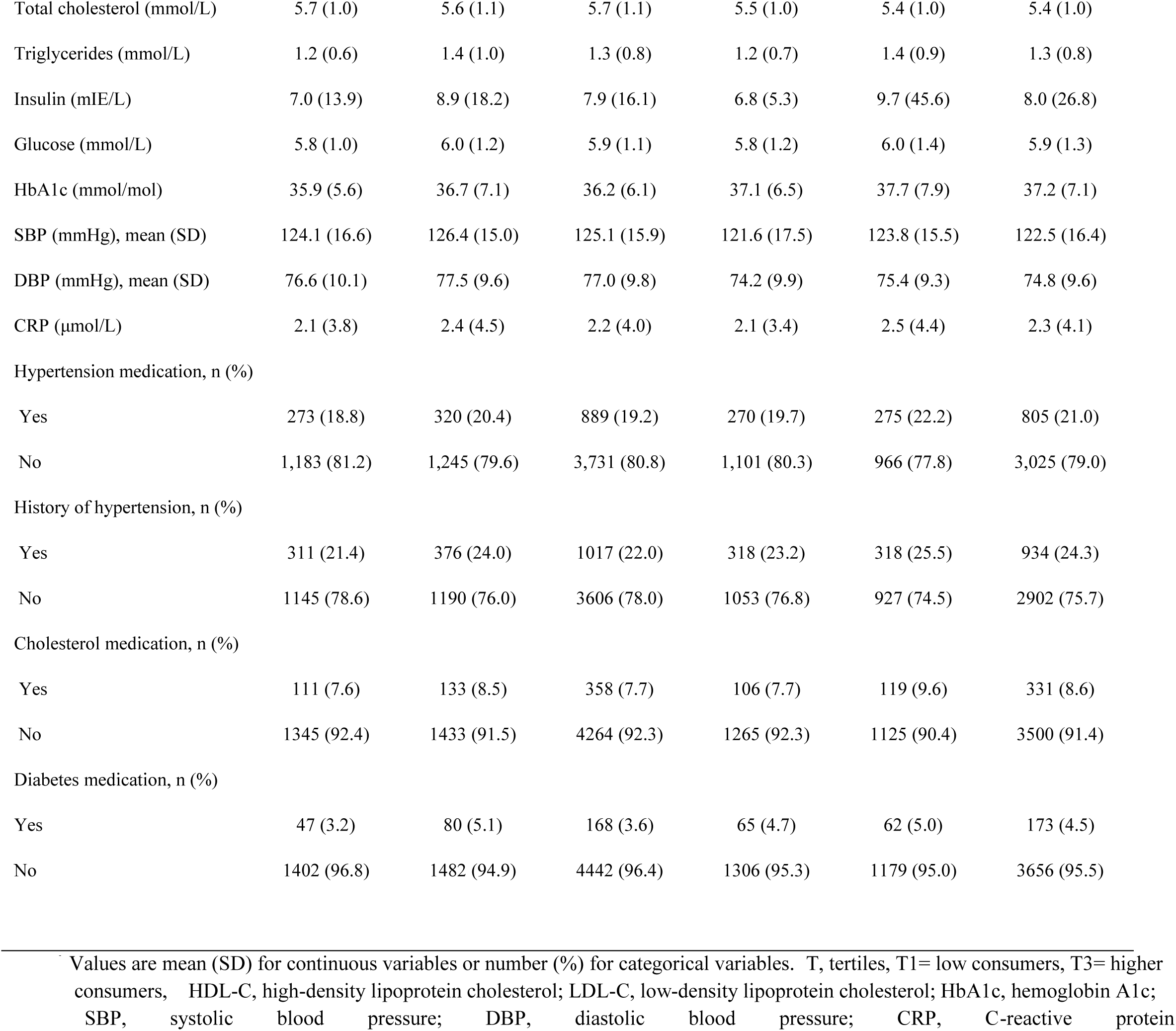
Characteristics of study participants based on the extreme tertiles (T1 and T3) of total meat intake.

### Metabolites associated with meat intake

We analyzed the association of white meat, unprocessed red meat, and processed red meat intake with plasma metabolites. In the basic model, 477 metabolites were associated with white meat **(Table S4)**, 489 with unprocessed red meat **(Table S5)**, and 386 with processed red meat intake **(Tables S6)**, respectively (*q*-value<0.05).

In the fully adjusted model, white meat intake was associated with 458 metabolites, including 130 positive and 328 negative associations **(Table S7)**. **Figure 1** shows the results for the top 20 metabolites associated with each meat type in the full model, selected based on the lowest *q*-values. Of the top findings, white meat intake demonstrated a strong positive association with 1-methyl-5-imidazolelactate (β=0.08; SE=0.003 per 20 grams of white meat intake; *q*-value=5.16×10^-78^) while the strongest negative association was observed with glutamine degradant (β=-0.03; SE=0.003 per 20 grams of white meat intake; *q-value*=4.05×10^-42^) **(Figure 1**, **Table S7)**.

**Figure 1:**
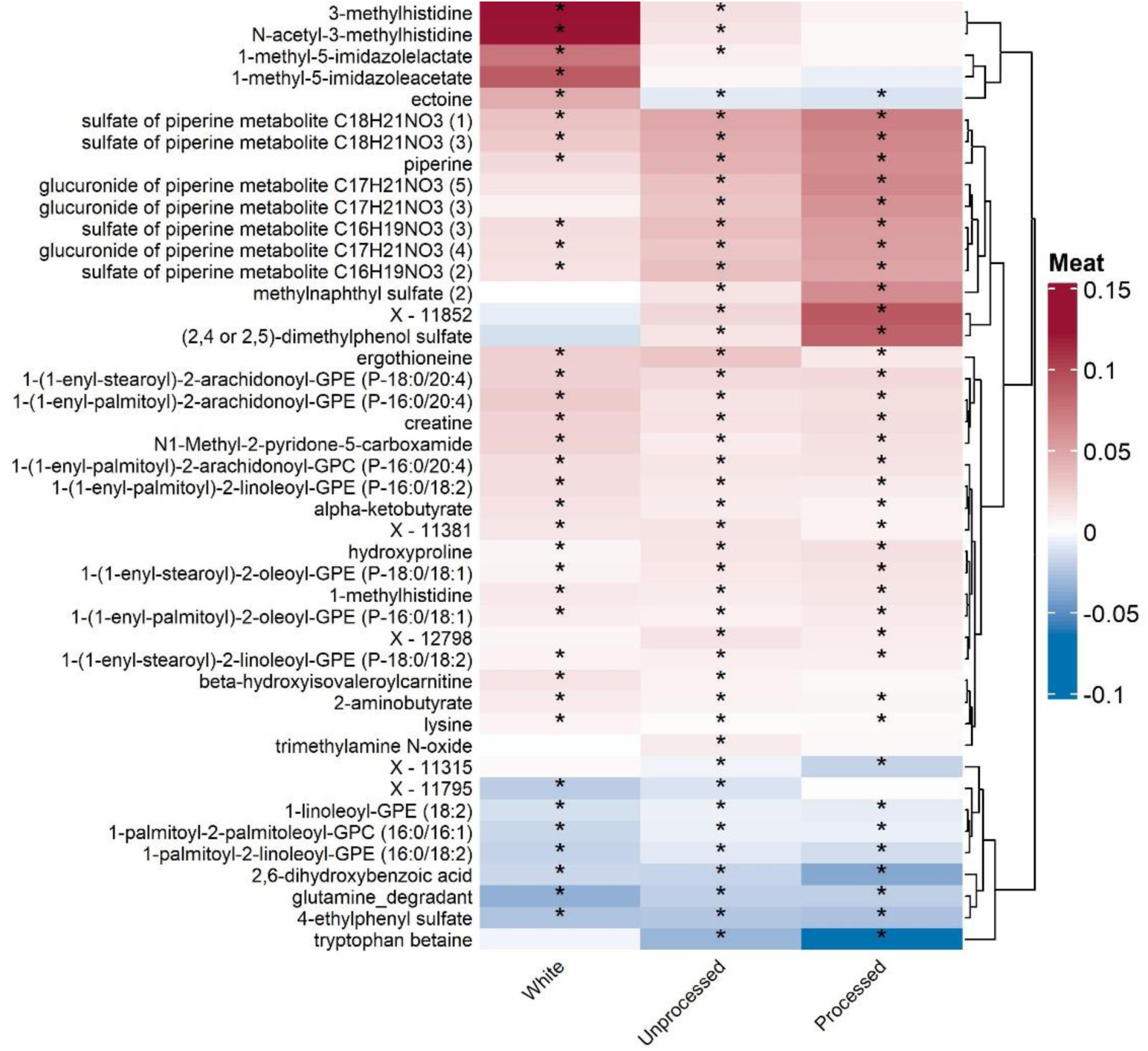
Heatmap illustrating the associations of meat intake with top 20 plasma metabolites. Linear regression coefficients (β) were adjusted for age, sex, batch effect, total energy intake, fiber intake, whole grain intake, fruit and vegetable intake, alcohol intake, physical activity, education, smoking history, hypertension medication, cholesterol medication, and diabetes medication. Asterisk (*) indicates statistical significance, as determined by false discovery rate–adjusted P values (q-values) <0.05.

In the fully adjusted model, unprocessed red meat intake was associated with 403 metabolites, comprising 165 positivel and 238 negative associations **(Table S8)**. Among the top findings, we observed that unprocessed red meat intake had a strong positive association with 1-(1-enyl-stearoyl)-2-arachidonoyl-GPE (P-18:0/20:4) (β=0.02; SE=0.001 per 20 grams of unprocessed red meat intake; *q-value*=1.64×10^-44^) while a strong negative association was observed for glutamine degradant metabolite (β=-0.02; SE=0.001 per 20 grams of unprocessed red meat intake; *q-value*=1.13×10^-33^), respectively **(Figure 1**, **Table S8)**. Similarly, processed red meat intake was associated with 368 metabolites, consisting of 168 positive and 200 negative associations **(Table S9)**. Strongest positive and negative associations were observed for the metabolites 1-(1-enyl-stearoyl)-2-arachidonoyl-GPE (P-18:0/20:4) (β=0.02; SE=0.001 per 20 grams of processed red meat intake; *q-value*=1.54×10^-33^) and 2,6-dihydroxybenzoic acid (β=-0.04; SE=0.003 per 20 grams of processed red meat intake; *q-value* =1.02×10^-25^), respectively (**Figure 1**, **Table S9)**.

**Figure 2** shows the plasma metabolites associated with the intake of white meat, unprocessed red meat, and processed red meat, categorized by metabolite classes. Metabolites positively associated with white meat intake primarily belonged to amino acids and lipids metabolite classes. However, for unprocessed red meat, the positively associated metabolites primarily belonged to lipids and xenobiotics, while for processed red meat they belonged to lipids, xenobiotics and uncharacterized molecules. **Figure S2** displays the specific and overlapping metabolites identified in association with meat exposures. There were 142 metabolites unique to white meat, 63 unique to unprocessed red meat, and 109 unique to processed red meat, while 145 metabolites overlapped among the three types.

**Figure 2.**
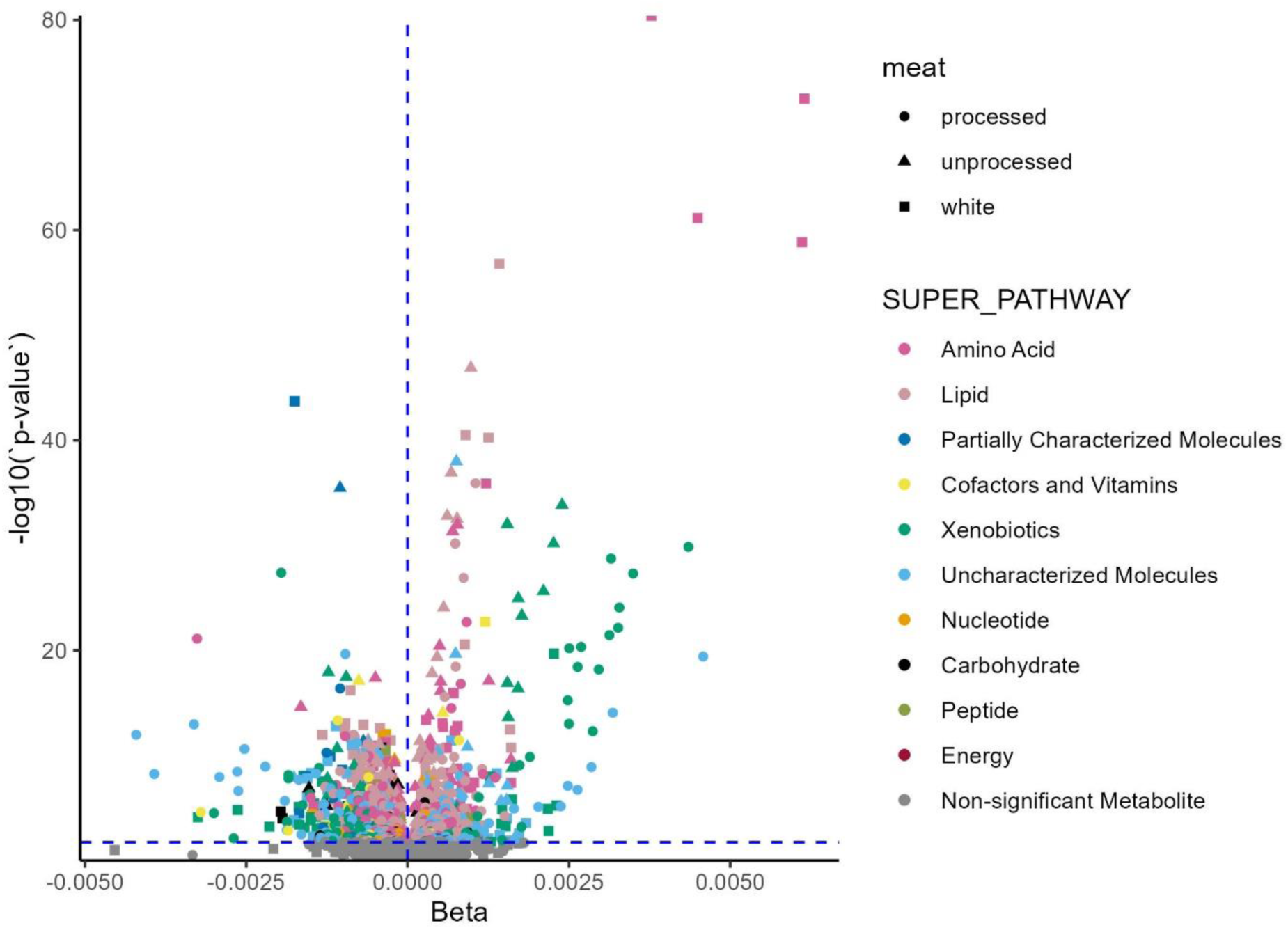
Volcano plot shows the association of dietary meat intake and plasma metabolites relative to their metabolite classes. Points above the red horizontal line indicate statistical significance. X-axis shows the meat-metabolites association beta coefficients while y-axis shows the -log10 *p*-values.

We also compared our findings with a recently published review article on metabolites associated with meat intake, which identified nine robust metabolites associated with different types of meat and reported them in both interventional and observational studies^34^ (**Figure S3, Table 2**). Similar to the previous findings, in the present study we observed white meat intake to be associated with 3-methylhistidine and carnitine, the latter of which was reported as O-acetyl-L-carnitine in the previous study. We also detected associations of both processed and unprocessed red meat with carnitine and carnosine. Furthermore, our analysis found that unprocessed red meat, but not processed red meat, was positively associated with trimethylamine *N*-oxide (TMAO).

**Table 2.** Meat intake in association with most frequently identified meat-associated metabolites using the SCAPIS-Uppsala and SCAPIS-Malmö cohorts. Analyses were adjusted for age, sex batch effect, total energy intake, fiber intake, whole grain intake, fruits and vegetables intake, alcohol intake, physical activity, education, smoking history of hypertension, medication (HTN, cholesterol, diabetes) and BMI. Asterisk (*) indicates statistically significant (q-value <0.05). The “+” sign indicates a positive association

|  |  | White meat |  |  |  | Unprocessed red meat |  |  | Processed red meat |  |  |
| --- | --- | --- | --- | --- | --- | --- | --- | --- | --- | --- | --- |
| Metabolites | Previous findings | n | Beta | SE | q-value | Beta | SE | q-value | Beta | SE | q-value |
| 3-methylhistidine | Chicken/Poultry (+) | 8193 | 0.12 | 0.01 | 1.9e-70* | 0.02 | 0.004 | 1.00E-04* | 0.01 | 0.01 | 4.1E-01 |
| Acetyl-carnitine (C2) | Poultry (+), Processed meat(+), red meat (+) | 8193 | -0.002 | 0.001 | 3.5E-01 | 0.003 | 0.001 | 6.0E-05* | 0.01 | 0.001 | 8.8E-05* |
| Carnitine | Red meat (+) | 8193 | 0.003 | 0.003 | 3.0E-03* | 0.005 | 0.001 | 5.3E-10* | 0.004 | 0.001 | 4.4E-03* |
| Carnosine | Chicken/Poultry (+), red meat (+) | 7771 | 0.01 | 0.01 | 9.4E-02 | 0.02 | 0.003 | 2.6E-07* | 0.02 | 0.005 | 1.7E-04* |
| Creatine | Total meat (+) | 8193 | 0.02 | 0.02 | 1.E-36* | 0.02 | 0.001 | 1.5E-30* | 0.02 | 0.002 | 2.9E-21* |
| Creatinine | Total meat (+) | 8193 | -0.0004 | 0.00 | 6.7E-01 | 0.000 | 0.000 | 3.4E-01 | 0.001 | 0.001 | 5.7E-01 |
| Glutamine | Total meat (+) | 7800 | 0.01 | 0.01 | 3.5E-01 | 0.01 | 0.007 | 3.9E-01 | 0.01 | 0.01 | 7.1E-01 |
| Trimethylamine N-oxide | Red meat (+) | 8193 | -0.001 | 0.003 | 9.1E-01 | 0.011 | 0.002 | 1.5E-07* | 0.004 | 0.003 | 4.0E-01 |
| Hydroxyproline | Total meat (+) | 8193 | 0.002 | 0.002 | 2.7E-02 | 0.01 | 0.001 | 5.8E-30* | 0.02 | 0.002 | 9.8E-16* |

### Pathway Enrichment Analysis of Metabolites Associated with Meat Intake

**Figure 3** and supplementary tables **(Tables S10-S18)**, depict the associations between meat-associated metabolites and their specific metabolic pathways. We observed positive enrichment of plasmalogen metabolic pathway for metabolites associated with the three meat types **(Tables S12**, **Table S15, Table S18)**. Moreover, white meat-associated metabolites were positively enriched for leucine, isoleucine, and valine metabolism (**Figure 3**, **Table S12**), while unprocessed red meat was enriched for histidine metabolism (**Table S15**). Metabolites associated with processed red meat showed positive enrichment in benzoate metabolism and fatty acid metabolism (acyl carnitine, medium chain) pathways (**Table S18)**.

**Figure 3:**
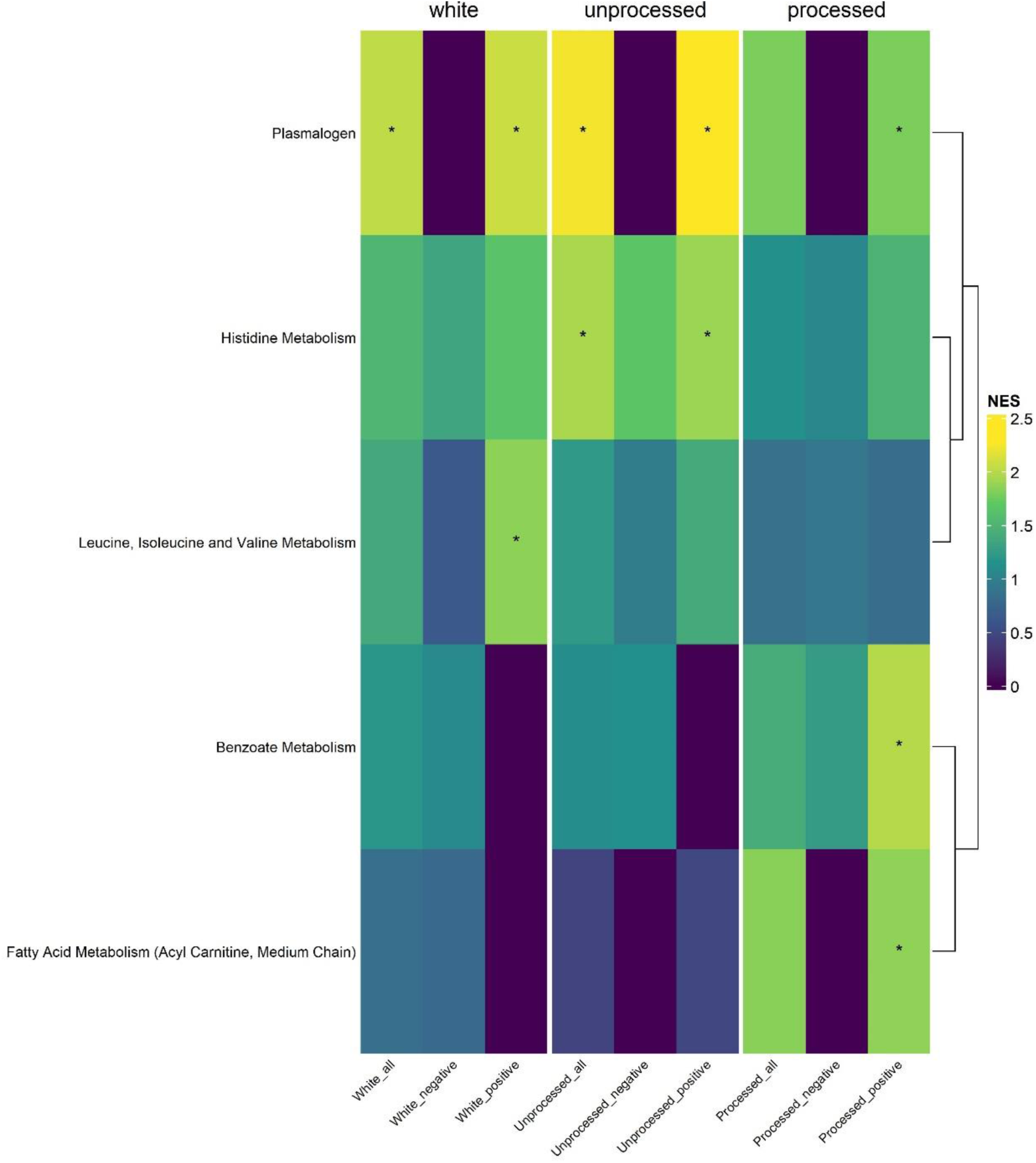
Gene enrichment analysis of meat-related metabolites. The heatmaps show the enriched metabolic sub-pathways in the associations between meat intake and plasma metabolites stratified by the direction of the associations. X-axis represents different meat types-plasma metabolites associations and y-axis represents different metabolic pathways. Black space represents missing pathways. NES, Normalized Enrichment Score

### Associations of meat-associated metabolites with cardiometabolic biomarkers, sublinical CVD markers and incident CVD

Next, we selected top 20 metabolites associated (based on *q*-values) with each meat type and analyzed their association with cardiometabolic biomarkers **(Table S19-S21)**, sublinical CVD markers (**Table S22-S24)** and incident CVD (**Table S25-S27)**.

We observed that metabolites positively associated with all three meat types were also positively related with ApoA1 and CRP **(Figure 4, Figure S3, Table S19-S21)**. In this pattern, three metabolites: 1-(1-enyl-stearoyl)-2-arachidonoyl-GPE (P-18:0/20:4), 1-(1-enyl-palmitoyl)-2-arachidonoyl-GPE (P-16:0/20:4), and 1-(1-enyl-palmitoyl)-2-arachidonoyl-GPC (P-16:0/20:4)—were positively related with IMT (**Figure 4**, **Table S22-S24**). We also observed that the metabolite 1-(1-enyl-palmitoyl)-2-arachidonoyl-GPC (P-16:0/20:4) was inversely related with IMGSM, EIDV, and EDV subclinical markers **(Figure 4, Table S22-S24**).

**Figure 4:**
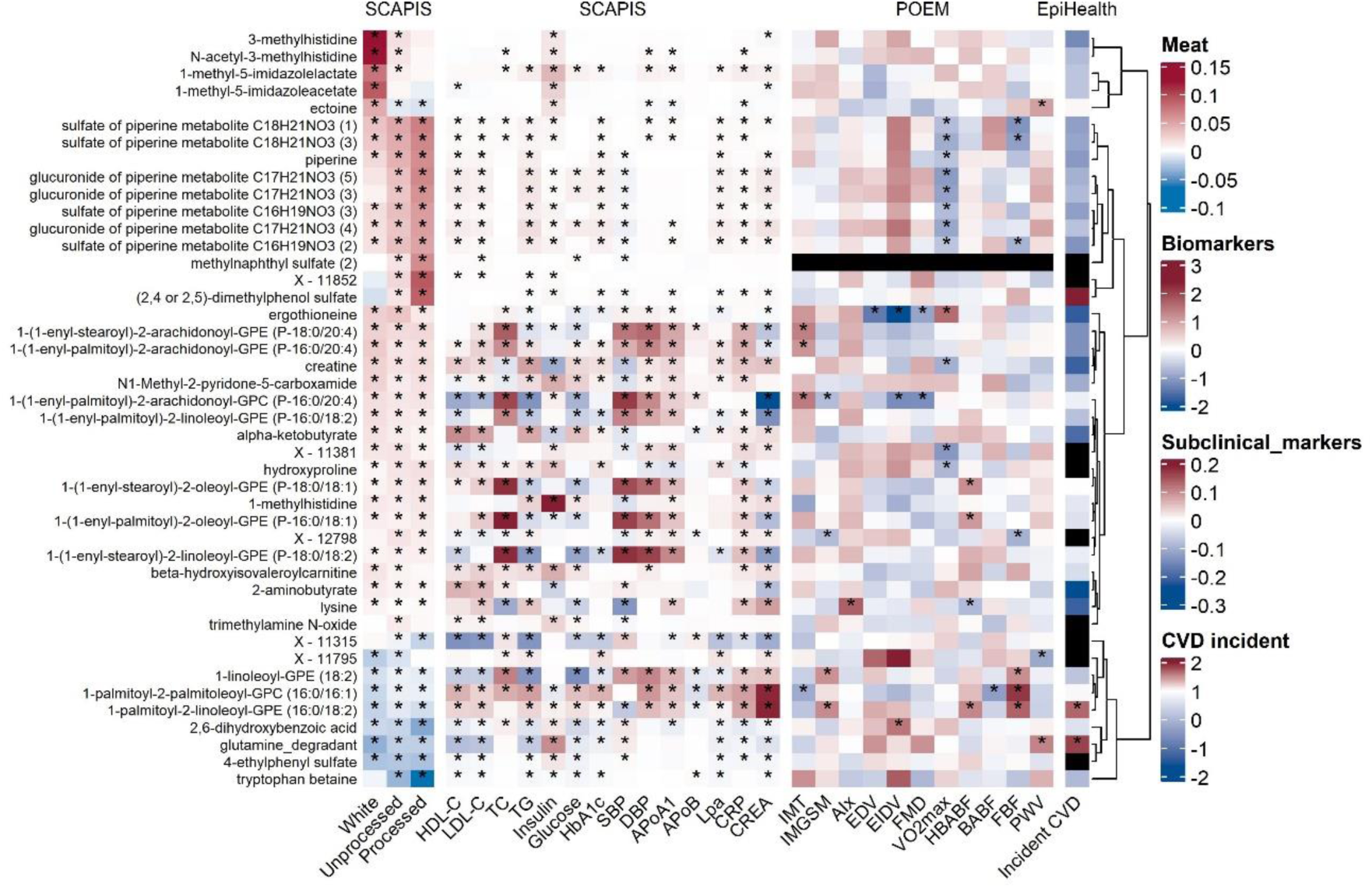
Heatmap illustrating the associations of the top 20 meat-related metabolites with cardiometabolic biomarkers in SCAPIS, subclinical markers in POEM study and incident CVD in EpiHealth. Cardiometabolic biomarkers and subclinical markers were scaled to the same units prior to statistical analysis. **Abbreviations**: HDL-C, high-density lipoprotein cholesterol; LDL-C, low-density lipoprotein cholesterol; TC, total cholesterol; TG, triglyceride; HbA1c, glycated hemoglobin; SBP, systolic blood pressure; DBP, diastolic blood pressure; ApoA1, apolipoprotein A1; ApoB, apolipoprotein B; Lpa, lipoprotein (a); CRP, C-reactive protein; CREA, creatinine, IMT, intima-media thickness of the carotid artery; IMGSM, intime-media grayscale median; AIx, Fasting Augmentation index; EDV, Endothelium dependent vasodilation; EIDV, Endothelium dependent vasodilation; FMD, flow-mediated vasodilation;VO2max; maximal oxygen consumption; HBABF, Hyperemia brachial artery blood flow; BABF, Resting brachial artery blood flow; FBF, Resting forearm blood flow; PWV, Pulse wave velocity; CVD, cardiovascular diseases. Asterisk (*) indicates statistically significant associations after 5% False Discovery Rate correction.

Moreover, we found that metabolites negatively associated with all three meat types were positively related with higher fasting insulin levels **(Figure 4, Figure S3, Table S19-S21)**. Two metabolites including 1-palmitoyl-2-linoleoyl-GPE (16:0/18:2) (HR: 1.32; 95% CI: 1.08, 1.62) and glutamine degradant (HR: 1.35; 95% CI: 1.07, 1.72) were related with an increased risk of incident CVD **(Figure 4, Table S25-S26)**.

We found that the metabolite ectoine, which was positively associated with white meat, but negatively associated with unprocessed and processed red meat, was related with higher PWV **(Figure 4, Table S22)**. Moreover, metabolites positively associated with processed red meat intake were also related with higher levels of insulin, HbA1c and Lpa **(Figure 4, Table S21),** and inversely related with VO2max **(Figure 4, Table S24).**

We observed that TMAO, metabolite that was positively associated with unprocessed red meat intake **(Figure 1)**, was related with higher levels of fasting insulin, HDL-C, LDL-C and glucose (**Figure 4, Table S20)**, but was not associated with subclinical CVD markers or incident CVD.

## Discussion

In this large population-based study, we investigated the association between white, unprocessed red, and processed red meat intakes and plasma metabolites. After adjusting for potential confounding factors, we identified 458 metabolites that were associated with white meat, 368 with unprocessed red meat, and 403 with processed red meat. Of these, 142 metabolites were unique to white meat, 63 to unprocessed red meat, and 109 to processed red meat, while 145 metabolites were overlapping among the three meat types. Metabolites associated with higher meat intake were linked to elevated levels of ApoA1 and CRP, while those specific to processed red meat were related with increased insulin, HbA1c, and Lpa levels. In an independent sample, several meat-related metabolites were associated with subclinical markers of CVD and incident CVD. For example, the metabolite ectoin, which is related to higher white meat intake, was associated with elevated PWV, while metabolites positively associated with processed red meat were related to VO2max. Our findings on meat-associated metabolites align with those from previously reported in a review article (**Table 2)**^34^. We observed similar associations between white meat and th 3-methylhistidine and carnitine. Red meat intake, including both unprocessed and processed types, was associated with carnitine and carnosine. However, we did not replicate the association between white meat and acetyl-carnitine (C2). In our study, acetyl-carnitine was inversely associated with white meat intake, whereas it was positively associated in the previous study^34^. Our study also revealed that several plasma metabolites were associated with white meat, unprocessed red meat, and processed red meat, which can be used for further validation and to expand upon previous studies in this area^10,12–19^.

Among the top 20 meat-associated metabolites, white meat intake was positively associated with 1-methyl-5-imidazolelactate, a product of histidine metabolism formed specifically through the methylation and subsequent decarboxylation of histidine derivatives^35^. This observation is likely due to the high protein content of white meat, which contains derivatives related to histidine metabolism^36,37^. Previous studies reported chicken/poultry intake to be associated with 3-methylhistidine, which is also derived from histidine metabolism^38,39^. The biological significance of 1-methyl-5-imidazoleacetate is less well characterized, and more research is needed to fully elucidate its role. Further, we found that the metabolite 1-(1-enyl-stearoyl)-2-arachidonoyl-GPE (P-18:0/20:4) was positively associated with both unprocessed and processed red meat. Our finding is consistent with the prior meat-metabolomic study in the EPIC-Norfolk cohort, which reported that red meat intake was positively associated with 1-(1-enyl-stearoyl)-2-arachidonoyl-GPE (P-18:0/20:4), however, red meat was not subclassified as unprocessed and processed in that study^20^.

Our study suggests that white meat and unprocessed red meat intake were negatively associated with glutamine degradants, indicating a potential role of meat intake in maintaining glutamine reserves. This finding indirectly suggests that vegeterians may have lower levels of specific amino acids, including glutamine precursors, even though they can synthesize glutamine from amino acids present in plant-based foods^40^. Additionally, our study shows that processed red meat was associated with lower plasma levels of 2,6-dihydroxybenzoic acid, though further research is needed, as the exact tole of this metabolite in human biology remains to be clarified.

In our study, we found that white meat-associated metabolites mainly consist of amino acids and lipid metabolites. It is biologically plausible that white meat contains generally higher levels of amino acids, such as glutamine compared to red meat^41^. The xenobiotics, such as the sulfate and glucuronide metabolites of piperine, were associated with both unprocessed and processed red meat. This association could be due to residual confounding or factors specific to meat processing, such as the type of additives and preservatives used in processed red meat, as well as other unmeasured variables. Wedekind *et al* reported that pepper alkaloids were associated with the intake of processed meat in one of the European EPIC cohorts^42^.

In the present study, the plasmalogen metabolic pathway was positively enriched for metabolites associated with all three meat types, indicating that these meat types were associated with an increase in metabolites involved in plasmalogen metabolism. Plasmalogens are vital for lipid raft and cholesterol-rich membrane stability, crucial for cellular signaling where meat, such as beef, pork, and chicken, is a primary dietary source^43^.

Moreover, our enrichment analysis revealed that the observed enrichment of white meat-associated metabolites and branched-chain amino acid metabolic pathways, such as histidine metabolism and leucine, isoleucine, and valine metabolism, is likely due to the direct intake of these metabolites from meat, rather than resulting from metabolic upregulation of these endogenous pathways. This limitation should be considered when interpreting the results, as distinguishing between exogenous and endogenous sources of these metabolites is challenging and requires further investigation. Our findings suggest that processed red meat metabolites were positively enriched for benzoate and medium chain acyl carnitine metabolism pathways^19^. This suggest that processed red meat may contain foreign chemical compounds that increase the risk of inflammation and metabolic abnormalities^6,19^.

We found that unprocessed red meat intake, but not processed red meat, was associated with TMAO. Similar to our findings, unprocessed red meat, but not processed meat, was associated with TMAO levels in a previous study ^3^. However, the literature on meat intake and TMAO is inconsistent. Previous studies have shown a positive association between red meat consumption and TMAO levels^3,44^. Moreover, TMAO, a metabolite formed by gut microbes, has often been associated with fish consumption^37,45^. These inconsistencies may arise from the overall influence of dietary patterns on the relationship between meat intake and TMAO levels, along with significant variations in meat consumption across different populations, study designs, and dietary habits. Furthermore, the finding that the association of plasma TMAO with higher levels of HDL cholesterol, LDL cholesterol, and glucose may indicate the implications of unprocessed red meat intake related TMAO for metabolic health and disease although causality cannot be inferred in our study. However, the lack of association between plasma TMAO levels and subclinical CVD markers and incident CVD, despite the positive association between elevated plasma levels of TMAO and CVD in previous findings is noteworthy^3,46^. We speculate that the inconsistent findings may be attributed to the small sample size used in POEM and EpiHealth.

Our study revealed specific patterns of meat-intake associated metabolites and their associations with cardiometabolic biomarkers, subclinical CVD markers and incident CVD. We found that metabolites positively associated with all three meat types, such as 1-(1-enyl-stearoyl)-2-arachidonoyl-GPE (P-18:0/20:4), were related to higher levels of ApoA1, CRP, and IMT, indicating a potential risk of CVD. Similary, metabolites negatively associated with all three meat types, such as 1-palmitoyl-2-palmitoleoyl-GPC (16:0/16:1) were associated with higher insulin levels. This association might result from complex metabolic or lifestyle factors, such as changes in lipid metabolism, affecting insulin sensitivity independently of dietary carbohydrate intake, which was relatively lower in the low meat intake group in our study^47^. Moreover, metabolites from processed red meat including sulfate and piperine were related with higher levels of insulin, HbA1c, Lpa, and lower values of VO2max, indicating impaired insulin sensitivity and reduced cardiovascular fitness in with higher intake.

In the present study, the finding that 1-palmitoyl-2-linoleoyl-GPE (16:0/18:2) and a glutamine degradant, which were inversely associated with all three meat types were associated with an increased risk of incident CVD. Lind *et al* reported that palmitoyl-oleoyl-GPE (16:0/18:1), another fatty acids metabolite integral to lipid metabolism was associated with incident CVD in the EpiHealth sample^48^. Our findings indicate that 3-methylhistidine, associated with white meat but not with processed red meat, shows a lower risk to CVD. Conversely, at the opposite end of the spectrum, dimethylphenol sulfate, associated with processed red meat but not white meat was related toincreased CVD risk. Additionally, ectoine, a metabolite associated with white meat but not processed red meat, was related to subclinical CVD markers such as a higher value of PWV; however, it was not associated with CVD risk. Although the results for meat associated metabolites and CVD risk may not be significant due to the limited number of cases in EpiHealth, these metabolites might serve as mediators of the effect of processed meat on CVD. Previous metabolomic studies have found that 3-methylhistidine was associated with future CVD risk^24,49^.

Our study has several strengths. First, a large-scale epidemiological sample enhances the robustness of the data and provides a comprehensive understanding of the associations between meat intake and metabolites. Second, participants were recruited using personal identification numbers, enabling randomized recruitment from the Swedish population register. Third, study participants were well-characterized with detailed phenotypic information, allowing for the control of potential confounders. Fourth, the classification of meat intake in this study may provide a specific metabolite association for different meat types, particularly specifying the metabolites associated with red meat types, as red meat intake is mostly explained for its association with adverse health outcomes, including CVD. However, several limitations need to be acknowledged. First, this is a cross-sectional study, and establishing a causal link between meat intake and plasma metabolites in SCAPIS would benefit from longitudinal study in the future. Second, participants are mainly from Sweden, aged 50–65, which affects generalizability and leaving the need to replicate our findings in different populations across other age groups. Third, the subjective nature of FFQ is imprecise and can be affected by systematic and random errors, such as underreporting of meat intake. Fourth, the relationships between meat intake and plasma metabolites may still be influenced by unmeasured confounders such as overall dietary habits. Fifth, even though a large number of metabolites were analyzed in our study, there may be metabolites that are related to meat intake that were not included in the assay.

## Conclusion

Our large-scale population-based study identified hundreds of plasma metabolites associated with self-reported white meat, unprocessed red meat, and processed red meat intake. Across all meat types, positively meat-associated metabolites were also related with cardiometabolic biomarkers such as higher plasma levels of ApoA1, CRP, and markers of subclinical CVD such as IMT. Processed red meat-associated metabolites were further associated with worse glycemic measures and reduced cardiovascular function. Two metabolites—1-palmitoyl-2-linoleoyl-GPE (16:0/18:2) and a glutamine degradant were associated with all three meat types and a higher risk of incident CVD. Our findings highlight plasma metabolites that may play a role in the association between meat intake and CVD, offering potential insights into the underlying mechanisms. However, further studies are needed to establish causality.

## Data Availability

The dataset supporting the findings of this research article was provided by the SCAPIS, EPIHEALTH, and POEM Data Access Board and are not publicly available due to confidentiality. Data can be shared upon reasonable request to the corresponding author, but only after obtaining permission from the Swedish Ethical Review Authority (https://etikprovningsmyndigheten.se)

## Acknowledgements

We acknowledge the financial support from the European Research Council [ERC-STG-2018-801965 (TF); ERC-CoG-2014-649021 (MO-M)], the Swedish Research Council [VR 2019-00977 (SCL), 2019-01471 (TF), 2018-02784 (MO-M), 2019-01015 (JA), 2020-00243 (JA), 2019-01236 (GE), 2022-01460 (SA)], the Swedish Heart-Lung Foundation [Hjärt-Lungfonden, 20230687 (TF), , 20200711 (MO-M), 20180343 (JA)], 20200173 (GE)], the Swedish Cancer Society [Cancerfonden, 2021 (SCL)],FORMAS [2020-00989 (SA)], Erik, Karin och Gösta Selanders Stiftelse [2020 (SA)], Åke Wibergs Stiftelse [2020 (SA)], Marcus Borgström Foundation [2020 (SA)], EFSD/Novo Nordisk [2020 (SA)], EpiHealth [2022 (SA)]. The main funding body of The Swedish CArdioPulmonary bioImage Study (SCAPIS) is the Swedish Heart and Lung Foundation. The study is also funded by the Knut and Alice Wallenberg Foundation, the Swedish Research Council, VINNOVA (Sweden’s Innovation agency), the University of Gothenburg and Sahlgrenska University Hospital, Karolinska Institutet and Region Stockholm, Linköping University and University Hospital, Lund University and Skåne University Hospital, Umeå University and University Hospital, Uppsala University and University Hospital. We would like to acknowledge the help of Biobank Sweden and the local biobank facilities for their services in handling of biological samples and biobanking. The computations and data handling were enabled by resources in project sens2019512 provided by the National Academic Infrastructure for Supercomputing in Sweden (NAISS) at Uppsala Multidisciplinary Center for Advanced Computational Science (UPPMAX), funded by the Swedish Research Council through grant agreement no. 2022-06725. The EpiHealth study is funded as a strategic research area by the Swedish government. POEM was funded by Uppsala University Hospital.

---

**Figure S1.**
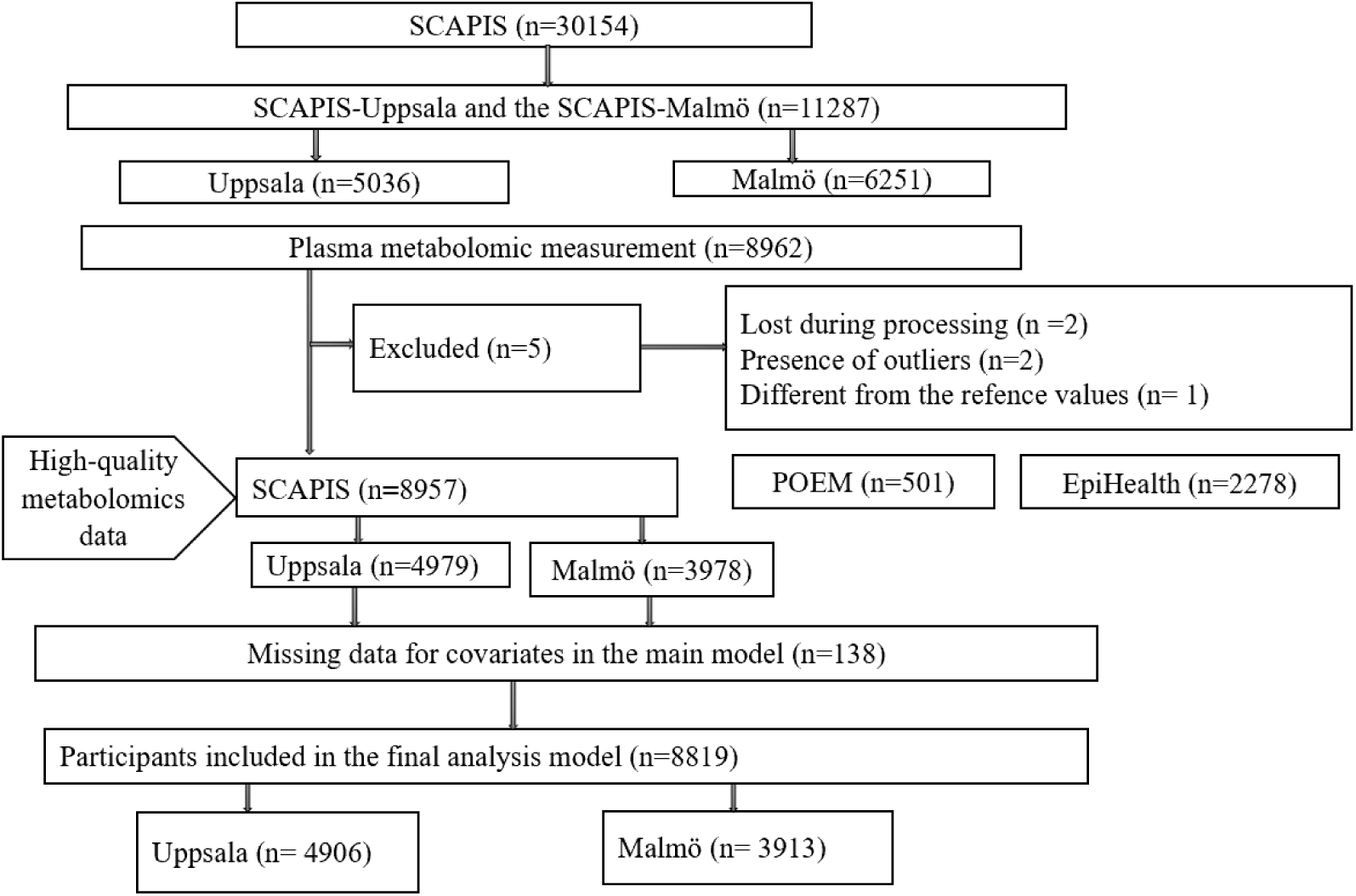
Flow chart of recruitment of participants to the meat-metabolome study.

**Figure S2:**
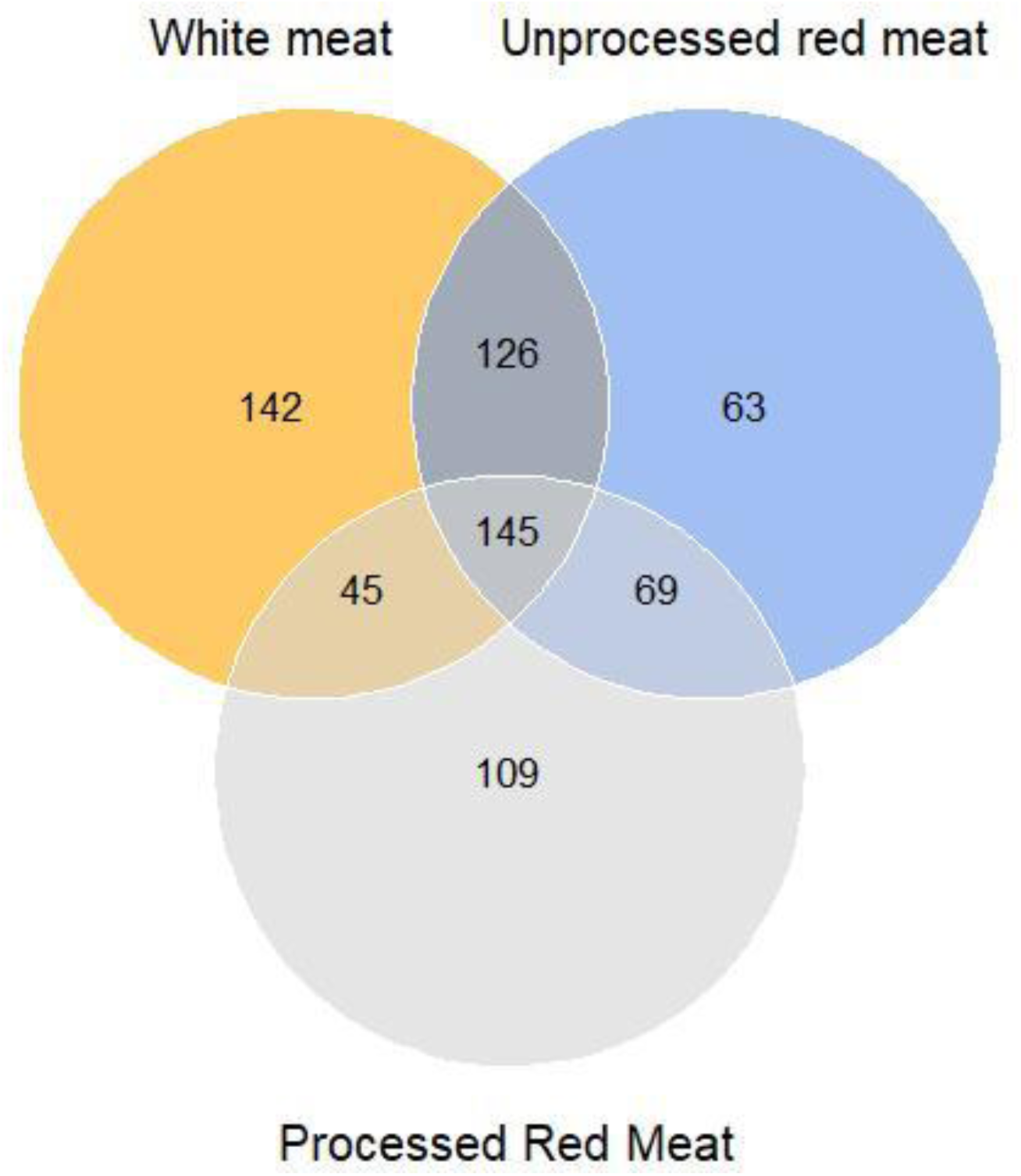
Venn diagram shows the specific and overlapping metabolites for the different types of meat (FDR <0.05)

**Figure S3.**
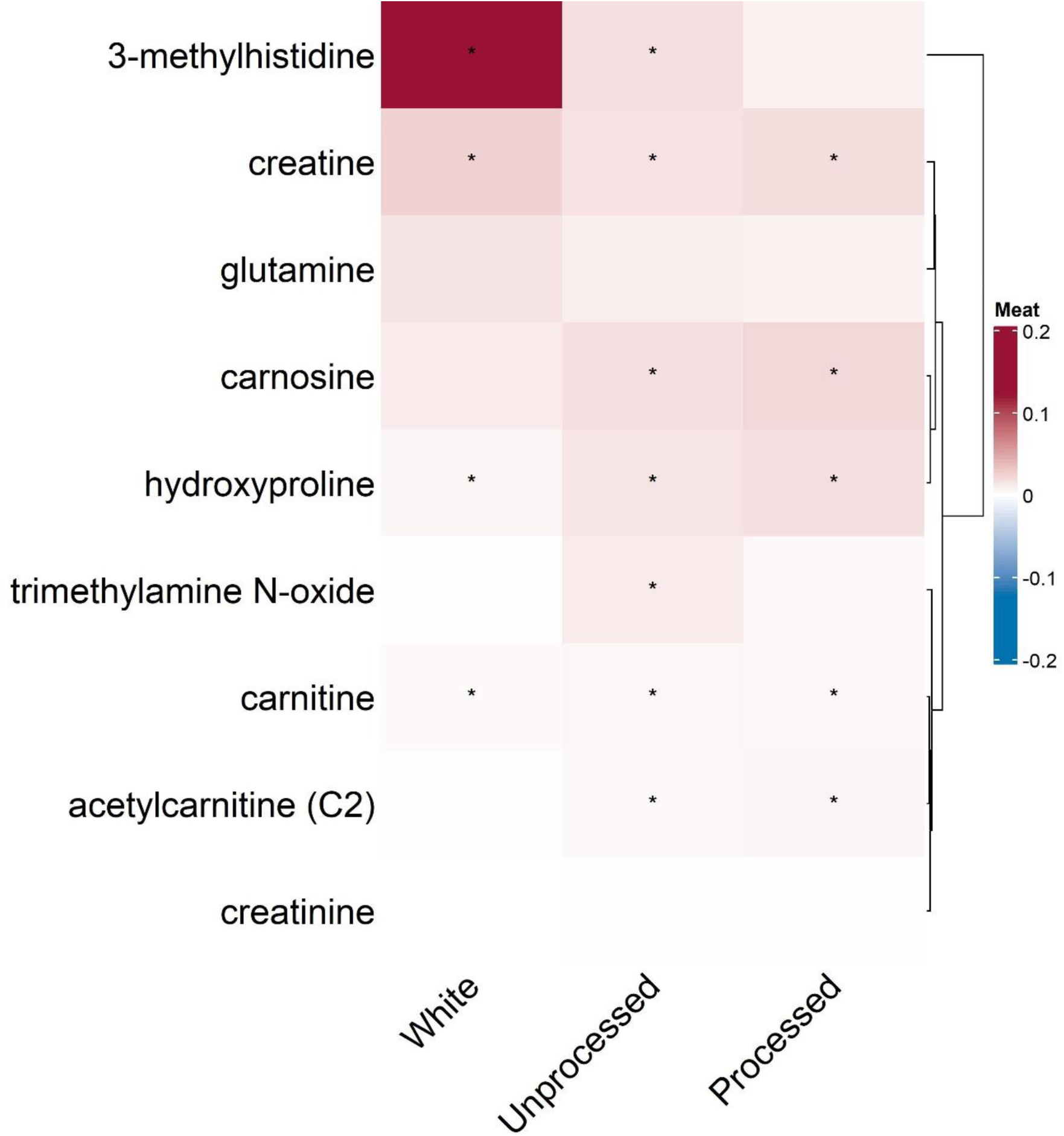
Heatmap illustrating the associations of meat intake with previously reported plasma metabolites. Asterisk (*) indicates statistically significant associations after 5% False Discovery Rate correction

**Figure S4:**
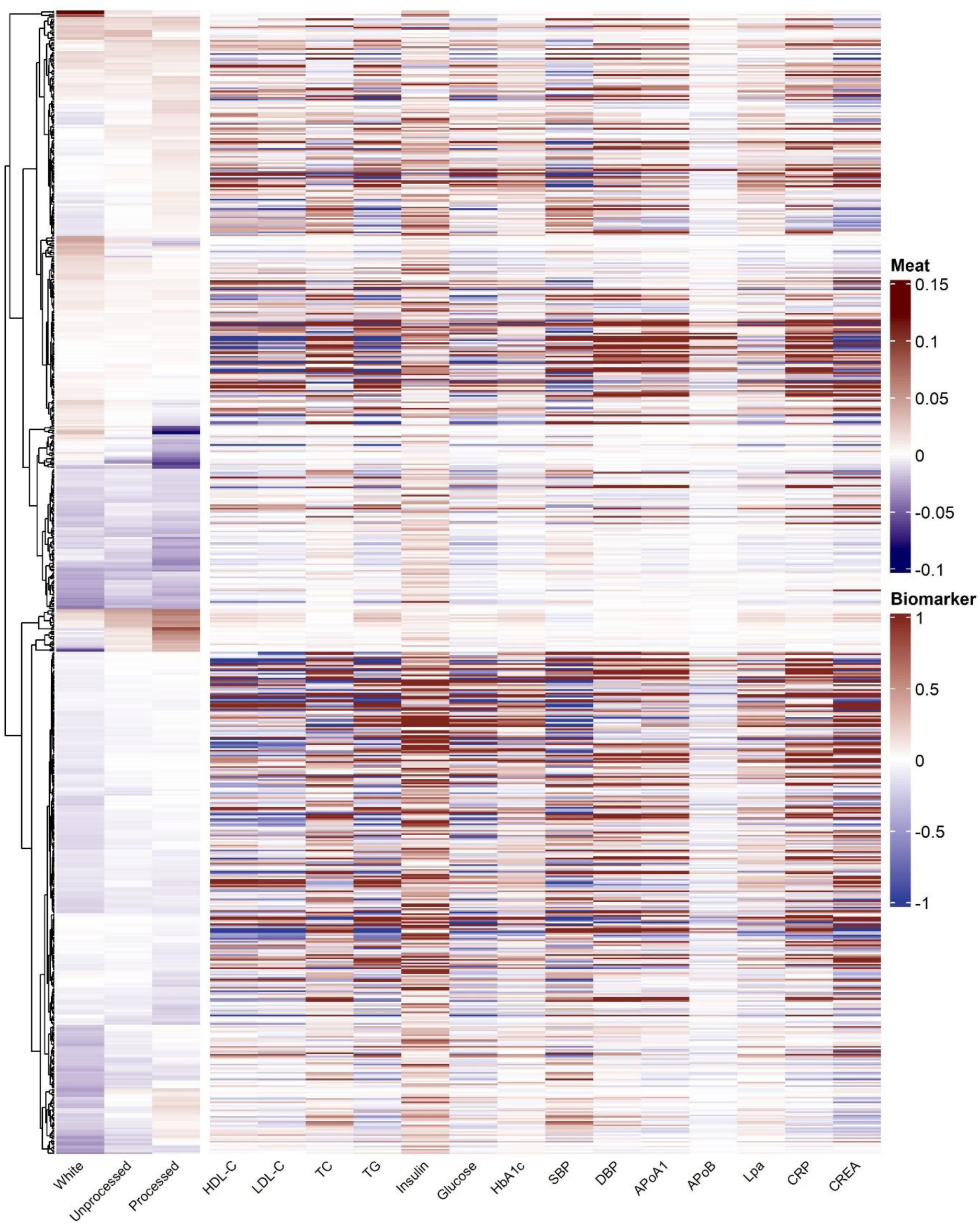
Heatmap illustrating the association of all meat-associated metabolites metabolites with cardiometabolic risk biomarkers (FDR <0.05). **Abbreviations**: HDL-C, high-density lipoprotein cholesterol; LDL-C, low-density lipoprotein cholesterol; TC, total cholesterol; TG, triglyceride; HbA1c, glycated hemoglobin; SBP, systolic blood pressure; DBP, diastolic blood pressure; ApoA1, apolipoprotein A1; ApoB, apolipoprotein B; Lpa, lipoprotein (a); CRP, C-reactive protein; CREA, creatinine

